# Inappropriate Dosing of Direct Oral Anticoagulants Among Very Elderly Inpatients With Atrial Fibrillation

**DOI:** 10.1101/2024.11.14.24317360

**Authors:** Ya-Tong Zhang, Jun-Peng Liu, Zi-Nan Zhao, Hong-Qiu Gu, Yi-Fan Na, Tian-Qi Zhang, Min Dong, Yu-Hao Wan, Min Zeng, Ning Sun, Cheng Wu, Jiefu Yang

**Author notes:** **Corresponding author:** Jun-Peng Liu, MD, PhD, Department of Cardiology, Beijing Hospital, National Center of Gerontology, Institute of Geriatric Medicine, Chinese Academy of Medical Sciences, No.1, Da Hua Road, Dongcheng District, Beijing 100730, China.

## Abstract

**Background:** Among very elderly patients with atrial fibrillation, the frequency of inappropriate direct oral anticoagulant (DOAC) dosing, associated factors, and temporal trends in practice are unknown.

**Objective:** To investigate the frequency of inappropriate DOAC dosing, associated factors, and temporal trends in very elderly patients with atrial fibrillation.

**Methods:** We retrospectively enrolled consecutive very elderly inpatients (≥80 years of age) diagnosed with atrial fibrillation at Beijing Hospital from January 2018 to August 2023, and discharged on DOACs were stratified according to receipt of underdosing, overdosing, or recommended dosing. Risk factors associated with underdosing or overdosing were identified using logistic regression. Cochran–Mantel–Haenszel analyses were used to assess temporal trends.

**Results:** We included 676 inpatients aged ≥80 years with atrial fibrillation (mean age 84.4±3.5 years, 53.1% female) prescribed a DOAC at hospital discharge (22.9% dabigatran, 62.3% rivaroxaban, 14.8% edoxaban). Recommended dosing occurred in 338 (50.6%) patients, underdosing in 308 (45.6%), and overdosing in 30 (4.4%). The overall rate of inappropriate dosing was 49.4%. Risk factors associated with underdosing included older age (OR = 1.98, 95% CI: 1.52–2.60, p < 0.001), lower CrCl (OR = 0.98, 95% CI: 0.97–0.99, p = 0.01), and non-internal medicine ward (OR = 2.15, 95% CI: 1.33–3.45, p = 0.002). The risk factor associated with overdosing was younger age (OR = 0.38, 95% CI: 0.19–0.75, p = 0.005). Over the study period, recommended dosing increased over time with a corresponding decline in inappropriate dosing, but these changes were not statistically significant.

**Conclusions:** Inappropriate DOAC dosing, especially underdosing, remains common in very elderly AF inpatients. This issue persists despite years passing, emphasizing the need for patient-focused, collaborative AF management and thorough prognostic studies.

**What Is New?:** Among AF patients aged 80 and above, nearly half experienced inappropriate NOAC dosing, with 92.3% of these cases being underdosed.

This situation has not shown significant improvement over the years.

Risk factors associated with underdosing included older age, compromised renal function, and hospitalization in non-internal medicine wards. Risk factor associated with overdosing was younger age.

**What Are the Clinical Implications?:** There is an urgent need for patient-centered, multidisciplinary AF management and shared decision-making, coupled with robust prognostic research focusing on very elderly AF patients.

## INTRODUCTION

Oral anticoagulant therapy has become the primary treatment for stroke prevention in atrial fibrillation (AF).^1–4^ In recent years, direct oral anticoagulants (DOACs) have gradually replaced warfarin as the preferred medication for anticoagulation therapy in non-valvular atrial fibrillation (NVAF).^2–4^ Compared to warfarin, DOACs offer advantages such as ease of use, no need for frequent INR monitoring, and fewer interactions with food and drugs, while also demonstrating good efficacy and safety in reducing the risk of stroke and major bleeding. The clinical effectiveness and safety of DOACs stem from appropriate dosing, which is based on guidelines and labeling, taking into account factors such as age, weight, kidney function, and concomitant medication use. However, substantial data indicate that inappropriate use of DOACs is significant and associated with increased risks—such as increased bleeding with DOAC overdosing and increased stroke risk with DOAC underdosing.^5, 6^

The inappropriate dosing of DOACs is particularly prevalent among elderly patients with AF, presenting a significant clinical concern. This issue stems from two primary factors. Firstly, the paucity of clinical evidence due to the exclusion of this demographic from most clinical trials has led to uncertainty regarding the safety and efficacy of currently recommended dosages. Secondly, anticoagulation therapy in elderly AF patients is inherently complex, compounded by aging process, including multimorbidity, kidney impairment, decreased drug metabolism, elevated fall risk, and increased risk of bleeding.^7, 8^

These challenges underscore the critical need for comprehensive studies focusing on DOAC use in the elderly AF population. To address this gap in knowledge, our investigation aims to elucidate the prevalence of inappropriate DOAC dosing, identify associated risk factors, and analyze temporal trends in this vulnerable patient cohort.

## METHODS

### Study Population

We conducted a retrospective chart review of all consecutive subjects aged 80 years and older with a diagnosis of AF who were discharged on a DOAC (rivaroxaban, dabigatran, or edoxaban) from Beijing Hospital between January 2018 and August 2023. Subjects were excluded from the study if they fulfilled any of the following criteria: 1) underwent left atrial appendage closure; 2) glomerular filtration rate <15ml/min/1.73m2; 3) incomplete data. This study was approved by the Ethics Committee of Beijing Hospital (approval no. 2023BYJYYEC-279-01). Demographic, clinical and nonclinical parameters were obtained by chart review.

Trained personnel abstracted clinical, demographic, laboratory, and treatment characteristics of these participants from the electronic medical record system. This included participants’ age, sex, ward of origin (internal medicine ward includes cardiology, respiratory, geriatric, neurology, rehabilitation, and traditional chinese medicine wards), comorbidities (such as diabetes mellitus, hypertension, heart failure, chronic kidney disease, and prior stroke), treatment variables (i.e., complete medication list), and laboratory values including serum creatinine and hemoglobin. History of bleeding (major bleeding or clinically relevant non-major bleeding), as defined by the International Society on Thrombosis and Hemostasis criteria,^9^ was also ascertained. Multimorbidity was defined as the number of comorbidities reported at baseline, when a patient presented at least 2 conditions.^10^ CrCl (creatinine clearance) was estimated with the Cockcroft-Gault formula. Polypharmacy was defined according to the number of drugs prescribed at baseline, as the presence of ≥7 different drugs taken by a patient.^11^ Antiplatelet drugs include aspirin, clopidogrel, and ticagrelor. Rhythm control drugs include propafenone, sotalol, and amiodarone. Rate control drugs include beta-receptor blockers, non-dihydropyridine calcium channel antagonists, and digoxin.

### Determining Appropriateness of DOAC Dose

Underdosing and overdosing were, respectively, defined as the administration of a lower or higher DOACs dose than recommended in the European Heart Rhythm Association (EHRA) consensus^12^ and package inserts. Inappropriate DOACs was defined as either underdosed or overdosed DOACs. For detailed recommended dosages, see Table S1.

### Statistical Analysis

Patients were divided into three groups: underdosing, overdosing, or recommended dosing of DOACs at discharge. We summarized continuous variables using means and standard deviations or medians and interquartile ranges, depending on the distribution’s normality. Categorical variables were summarized by count and percentages. We used analysis of variances or Kruskal– Wallis for continuous variables and the chi-square test for categorical variables to compare between groups.

To identify the risk factors of underdosing or overdosing, we conducted backward selection of logistic regression. The candidate risk factors, including age, sex, BMI (body mass index), cardiology ward, internal medicine ward, Praxysmal AF, CHA_2_DS_2_-VASc score, HAS-BLED score, multimorbidity, history of bleeding, dementia, ALT (alanine aminotransferase), AST (aspartate aminotransferas), Hb (hemoglobin), CrCI, polypharmacy, antiplatelet drug, rhythm control drug, rate control drug, NSAID (non-steroidal anti-inflammatory drug) were determined through a comprehensive consideration of the literature review, clinical experience, and data imbalance. We used Cochran–Mantel–Haenszel tests to assess the calendar trends of underdosing or overdosing.

## RESULTS

Between January 2018 and August 2023, a total of 676 consecutive elderly AF patients aged ≥80 years receiving DOAC treatment were studied (Figure 1). The mean age was 84.4±3.5 years, 53.1% were female, and the mean body mass index was 24.6±3.7 kg/m2. 60.4% had paroxysmal AF, and the mean CHA_2_DS_2_-VASc score and HAS-BLED score were 5.0±1.5 and 1.6±0.7, respectively. Figure 2 illustrates the distribution of underdosing, recommended dosing, and overdosing by DOAC type.

**Figure 1.**
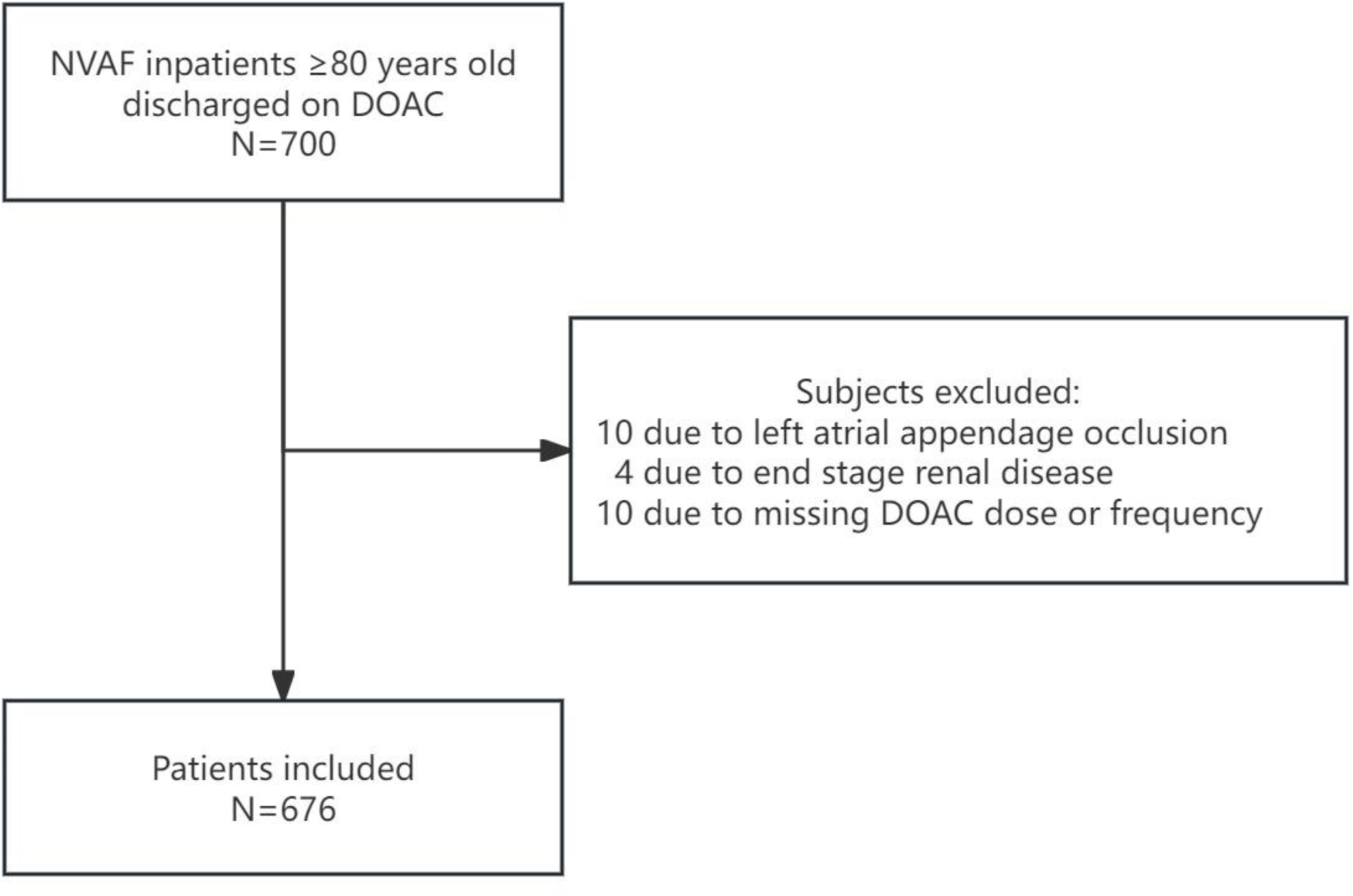
Flowchart of patient enrollment.

**Figure 2.**
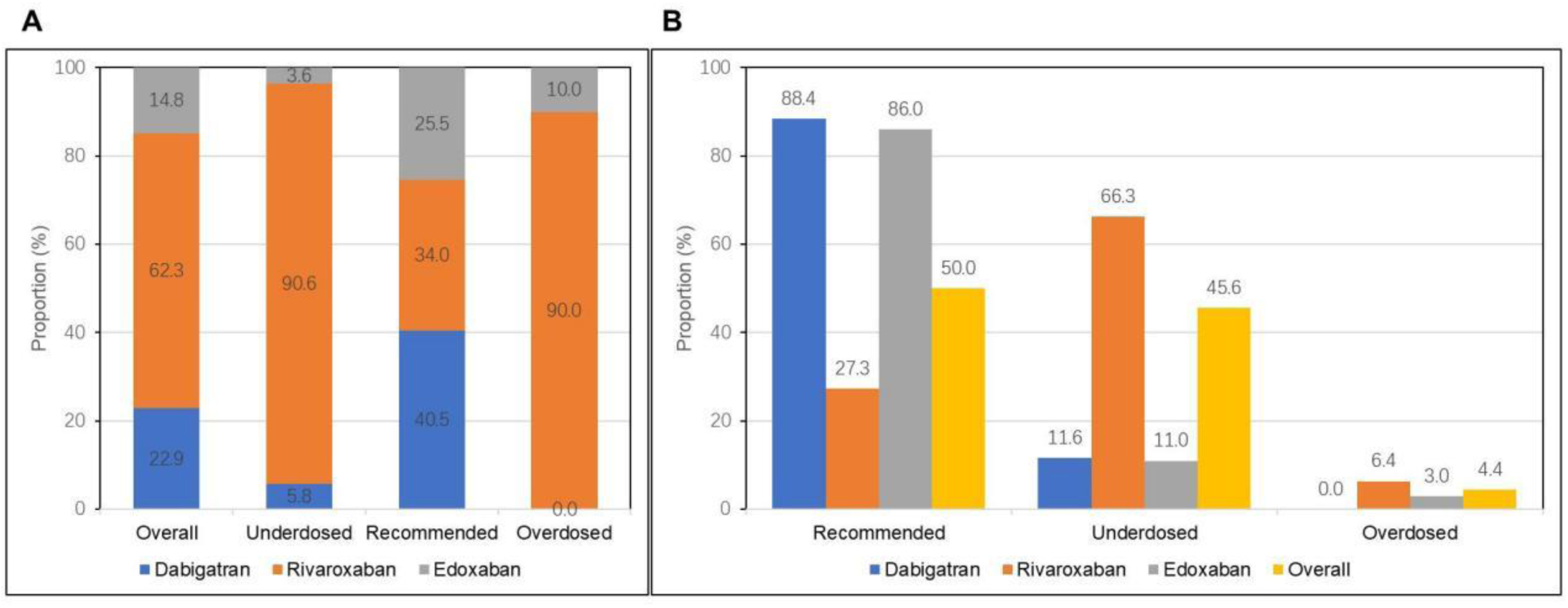
Frequency of NOAC prescription. (A) frequency of overall, underdosing, overdosing, and recommended dosing by each NOAC type; (B) frequency of NOAC dosing by recommended dosing, underdosed, or overdosed.

308 patients (45.6%) received a DOAC dose lower than recommended, 338 (50.6%) received the recommended DOAC dose, and 30 (4.4%) received a DOAC dose higher than recommended (Table 1, Table S2 and S3). Compared to those discharged on recommended DOAC dosing, patients who received underdosed DOACs were older (85.4±3.7 versus 83.6±3.2 years), had lower BMI (24.2±4.0 versus 25.1±3.5), higher HAS-BLED score (1.7±0.7 versus 1.5±0.7), were less frequently found in Cardiology ward (44.5% versus 59.8%) and Internal Medicine ward (77.9% versus 88.8%), were more bedridden (17.2% versus 8.9%), more frequently had a prior hemorrhage (8.4% versus 2.7%), lower Hb (hemoglobin) (119.7±17.8 versus 124.4±16.2 g/L) and CrCl (47.9±15.9 versus 53.9±16.0 ml/min). Relative to those discharged on recommended DOAC doses, patients who received overdosed DOACs were younger (82.7±2.4 versus 83.6± 3.2 years), less frequently found in the Cardiology ward (30.0% versus 59.8%), and more likely to have a prior hemorrhage (13.3% versus 2.7%). They also had higher CrCl (58.9±19.0 versus 53.9 ±16.0 ml/min) and lower Alb (albumin) (35.9±5.6 versus 37.4±3.8 g/L).

**Table 1.** Baseline patient characteristics.

|  | Underdosed<br>n=308 (45.6%) | Recommended<br>n=338 (50.6%) | Overdosed<br>n=30 (4.4%) | p Value |
| --- | --- | --- | --- | --- |
| Age, y | 85.3±3.7 | 83.6±3.2 | 82.7±2.4 | **<0.001** |
| Male | 135 (43.8) | 165 (48.8) | 17 (56.7) | 0.25 |
| Smoking | 104 (33.8) | 92 (27.2) | 9 (30.0) | 0.19 |
| Drinking | 102 (33.1) | 93 (27.5) | 8 (26.7) | 0.28 |
| BMI, kg/m<sup>2</sup> | 24.2±4.0 | 25.1±3.5 | 24.2±3.1 | **0.01** |
| Cardiology Ward | 137 (44.5) | 202 (59.8) | 9 (30.0) | **<0.001** |
| Internal Medicine Ward | 240 (77.9) | 300 (88.8) | 22 (73.3) | **<0.001** |
| Bedridden | 53 (17.2) | 30 (8.9) | 5 (16.7) | **0.006** |
| Praxysmal AF | 194 (63.0) | 196 (58.0) | 18 (60.0) | 0.43 |
| CHA2DS2-VASc score | 5.1±1.4 | 5.0±1.5 | 4.8±1.9 | 0.41 |
| HAS-BLED score | 1.7±0.7 | 1.5±0.7 | 1.6±0.7 | **0.04** |
| Multimorbidity | 301 (97.7) | 327 (96.7) | 27 (90.0) | 0.06 |
| DVT | 31 (10.1) | 15 (4.4) | 6 (20.0) | **<0.001** |
| PE | 24 (7.8) | 7 (2.1) | 6 (20.1) | **<0.001** |
| Surgical<br>thromboprophylaxis | 10 (3.2) | 4 (1.2) | 3 (10.0) | **0.007** |
| HTN | 251 (81.5) | 265 (78.4) | 21 (70.0) | 0.27 |
| CAD | 151 (49.0) | 162 (47.9) | 10 (33.3) | 0.26 |
| MI | 40 (13.0) | 31 (9.2) | 4 (13.3) | 0.28 |
| HF | 60 (19.5) | 62 (18.3) | 4 (13.3) | 0.70 |
| CIED | 56 (18.2) | 87 (25.7) | 2 (6.7) | **0.009** |
| catheter ablation | 45 (14.6) | 89 (26.3) | 8 (26.7) | **<0.001** |
| Diabetes | 114 (37.0) | 124 (36.7) | 8 (26.7) | 0.52 |
| Stroke/TIA | 97 (31.5) | 117 (34.6) | 10 (33.3) | 0.70 |
| SE | 10 (3.2) | 11 (3.3) | 2 (6.7) | 0.60 |
| PVD | 84 (27.3) | 72 (21.3) | 11 (36.7) | 0.06 |
| OSAS | 9 (2.9) | 15 (4.4) | 0 (0.0) | 0.33 |
| CKD | 40 (13.0) | 30 (8.9) | 4 (13.3) | 0.23 |
| COPD | 24 (7.8) | 16 (4.7) | 1 (3.3) | 0.22 |
| History of bleeding | 26 (8.4) | 9 (2.7) | 4 (13.3) | **0.001** |
| Dementia | 16 (5.2) | 9 (2.7) | 0 (0.0) | 0.13 |
| Hb g/L | 119.7±17.8 | 124.4±16.2 | 120.3±24.0 | **0.003** |
| ALT U/L | 14.0 (10.0,22.0) | 15.0 (11.0,21.0) | 14.5 (10.0,27.0) | 0.71 |
| AST U/L | 19.5 (16.0,25.0) | 19.0 (16.0,24.0) | 19.0 (16.0,24.0) | 0.79 |
| Scr umol/L | 90.1±32.3 | 86.2±34.7 | 80.6±28.5 | 0.08 |
| CrCl, ml/min | 47.9±15.9 | 53.9±16.0 | 58.9±19.0 | <0.001 |
| Alb, g/l | 36.1±3.9 | 37.4±3.8 | 35.9±5.6 | <0.001 |
| D-dimer, ng/ml | 273.0 (130.5, 636.0) | 188.0 (105.0, 365.5) | 475.0 (196.0, 1625.0) | 0.44 |
| Polypharmacy | 183 (59.4) | 210 (62.1) | 23 (76.7) | 0.17 |
| Antiplatelet drug | 44 (14.3) | 38 (11.2) | 4 (13.3) | 0.51 |
| Rhythm control drug | 58 (18.8) | 69 (20.4) | 2 (6.7) | 0.18 |
| Rate control drug | 155 (50.3) | 195 (57.7) | 19 (63.3) | 0.11 |
| NSAID | 6 (1.9) | 2 (0.6) | 1 (3.3) | 0.20 |

**Note:** Values are presented as mean ± SD or median (interquartile range: 25th to 75th percentiles) or number (percentage).

At multivariate logistic regression analysis (Table 2 and 3), older age (OR = 1.98, 95% CI: 1.52– 2.60, p < 0.001), lower CrCl (OR = 0.98, 95% CI: 0.97–0.99, p = 0.01), and non-internal medicine ward (OR = 2.15, 95% CI: 1.33–3.45, p = 0.002) were independently associated with underdose prescription. Younger age (OR = 0.38, 95% CI: 0.19 – 0.75, p = 0.005) was independently associated with overdose prescription.

**Table 2.** Risk factors associated with Underdosing or Overdosing.

| Factor | Unadjusted Analysis |  |  | Adjusted Analysis |  |  |
| --- | --- | --- | --- | --- | --- | --- |
|  | *OR* | *95%CI* | *P value* | *OR* | *95%CI* | *P value* |
| Risk factor of underdosing |  |  |  |  |  |  |
| Age | 2.22 | 1.74-2.82 | <0.001 | 1.98 | 1.52-2.60 | <0.001 |
| CrCl | 0.98 | 0.97-0.99 | <0.001 | 0.98 | 0.97-0.99 | 0.001 |
| Non-internal medicine ward | 2.09 | 1.39-3.17 | <0.001 | 2.15 | 1.33-3.45 | 0.002 |
| Risk factor of Overdosing |  |  |  |  |  |  |
| Age | 0.40 | 0.20-0.80 | <0.010 | 0.38 | 0.19-0.75 | 0.005 |

**Abbreviations:** CrCl=creatinine clearance

Figure 3 shows temporal trends in the rate of recommended dosing, underdosing, and overdosing of DOACs. Although there were fluctuations and the differences are not significant, it can be observed that the proportion of recommended dosing (51.2% in 2018 to 58.8% in 2023, p=0.09) shows an increasing trend, while the proportions of underdosing (42.7% in 2018 to 39.5% in 2023, p=0.19) and overdosing (6.1% in 2018 to 1.7% in 2023, p=0.32) show a gradually decreasing trend.

**Figure 3.**
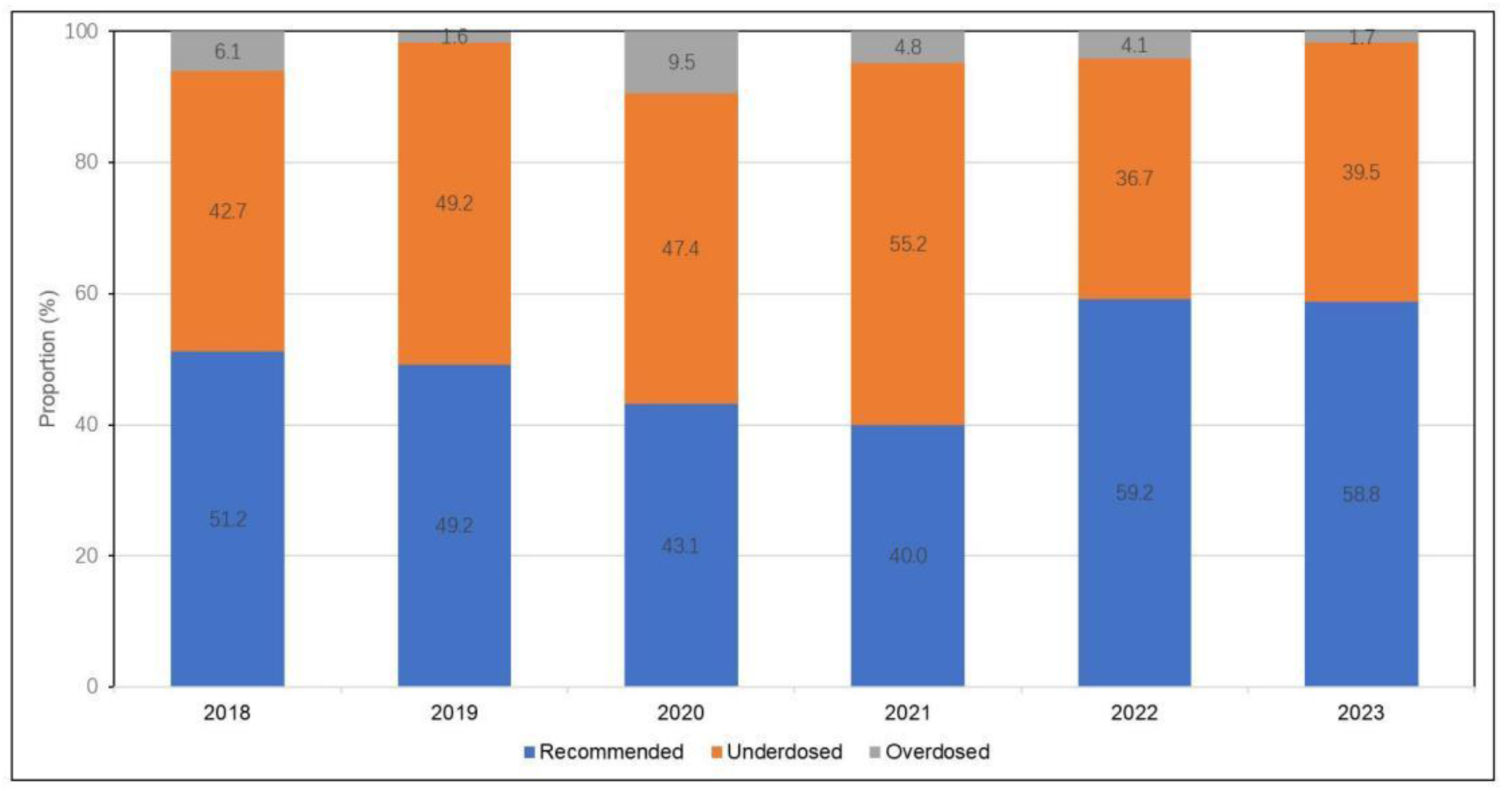
Trends in the rates of recommended, underdosing, and overdosing of DOAC.

## DISCUSSION

The main finding of this study are: (1) Inappropriate dosing of DOACs was relatively common among AF inpatients aged ≥80 years, with an overall inappropriate dosing rate of 49.4%. (2) Underdosing (45.6%) was more common than overdosing (4.4%). (3) Risk factors associated with underdosing included: older age, compromised renal function (decreased creatinine clearance), and hospitalization in non-internal medicine wards. (4) The main risk factor associated with overdosing was younger age. (5) Although there was no statistically significant difference, from 2018 to 2023, the use of recommended dosages showed an increasing trend year by year (from 51.2% to 58.8%), while the proportions of underdosing and overdosing showed a gradual decreasing trend (from 42.7% to 39.5%, and from 6.1% to 1.7%, respectively).

With global population aging, especially in China, stroke prevention in elderly patients, particularly those of advanced age with AF, faces unprecedented challenges. More worryingly, the issue of inappropriate DOAC dosing is particularly prominent in the very elderly AF population. Our survey results showed that the proportion approaches half, and other studies are not optimistic. A study from Italyshowed that among AF patients aged 80 and above, the rate of inappropriate DOAC use was 29%, with 74.4% being underdosed and 25.6% being overdosed.^13^ A German study found that in AF patients aged 85 and above, the rate of inappropriate DOAC use was 26.1%, all of which were underdosed.^14^ Data from a French medical center indicated that for AF patients aged 80 and above, the rate of inappropriate DOAC use was 40.0%, with 88.0% being underdosed.^15^ Another survey of elderly hospitalized patients with an average age of 82±8 years showed that the rate of inappropriate DOAC use was as high as 54.4%, of which 93.8% were underdosed.^16^ These research results indicated that underdosing of DOAC is the most prominent issue among inappropriate DOAC dosing in very elderly AF populations. Our research findings were consistent with this, with DOAC underdosing accounting for up to 90.7% of cases. Very elderly AF patients receiving low-dose DOAC typically have the following characteristics: older age, lower BMI or body weight, higher bleeding risk, and poorer renal function. Multiple studies also supported these findings^13, 15^. Additionally, we analyzed the differences between various departments and found that DOAC underdosing was less common in cardiology and internal medicine wards. Interestingly, this contrasts with a previous French study, which found underdosing to be more prevalent in cardiology wards.^15^ This phenomenon may reflect differences in experience and education levels regarding stroke prevention in AF across different departments and healthcare systems.

Previous studies^5, 17, 18^, including younger AF cohorts, had confirmed that advanced age, renal dysfunction, weight, and bleeding risk were independent risk factors for underdosing of DOACs. However, research on very elderly AF populations was relatively limited. Carbone et al.^13^ found that in very elderly AF patients, male gender, coronary heart disease, and low BMI were independently associated with underdosing of DOACs.Cavillon Decaestecker et al.^15^ reported that neurology wards were negatively correlated with underdosing of DOACs, while the use of antidepressants was positively correlated. Our research analysis also confirmed that in very elderly AF patients, advanced age and renal dysfunction were independently associated with underdosing of DOACs. Additionally, we explored the correlation between hospital wards and underdosing of DOACs. Similarly, to the analysis by Cavillon Decaestecker et al.,^15^ our analysis identified internal medicine wards (including neurology) were negatively correlated with underdosing. This finding further emphasized the importance of multidisciplinary management for AF. On the other hand, previous studies and our research had shown that although the proportion was low, there was still a problem of DOAC overdosing in very elderly AF patients. Carbone et al.’s study had shown that diabetes and history of bleeding were positively correlated with overdosage, while age and BMI were negatively correlated.^13^ Our study had also confirmed an independent correlation between age and DOAC overdosing.

It was worth noting that our study found that the appropriateness of DOAC use in very elderly AF patients had improved year by year, although this improvement was not significant. In contrast, the situation was more optimistic for younger AF patient groups, where the appropriate use of DOACs showed significant annual improvement. The reasons for this difference, besides the complex characteristics of the very elderly population itself, also lay in the lack of evidence-based medical evidence for this group. On one hand, patients aged 80 and above are typically excluded from clinical trials of DOACs^2–4^. In previous landmark clinical trials of DOACs, the mean or median age of participants ranged from 70 to 73 years.^2–4^ Therefore, the clinical guidelines recommending the use of DOACs based on these trials may not be entirely applicable to very elderly patients. On the other hand, there is still a lack of high-quality research on underdosing than recommended dosing of DOACs. The ELDERCARE-AF (Edoxaban Low-Dose for Elder Care Atrial Fibrillation Patients) trial^19^ is the only randomized controlled trial to date that has studied underdosing of NOACs in AF patients aged 80 and above. This study demonstrated that a once-daily 15 mg dose of edoxaban was superior to placebo in preventing stroke or systemic embolism and did not result in a significantly higher incidence of major bleeding compared to placebo. However, evidence comparing underdosing than recommended dosing of DOAC primarily comes from observational cohort studies, which involved relatively younger subjects and have inconsistent conclusions. The GARFIELD-AF (Global Anticoagulant Registry in the FIELD-AF) study^17^, which included patients with an average age of 72 years, revealed in its 2-year follow-up data that underdosing of DOACs was associated with an increased risk of all-cause mortality. Interestingly, despite a significantly reduced bleeding risk, there were no significant differences in the risks of stroke, systemic embolism, or major bleeding compared to recommended doses. Similarly, data from the ORBIT-AF II registry study, focusing on AF patients with an average age of 71 years, showed that patients receiving underdosed DOACs had a significantly higher rate of cardiovascular hospitalization within one year.^5^ The two-year follow-up data further indicated an increase in thromboembolic events and mortality among these patients.^20^ While these differences weren’t statistically significant after adjusting for variables, the overall data trend still leaned towards supporting the use of recommended DOAC doses. Meanwhile, multiple studies from different regions with comparable age groups of study populations have also confirmed that underdosing of DOACs not only fails to reduce the risk of bleeding, systemic embolism, or all-cause mortality in atrial fibrillation patients, but may also significantly increase the risk of stroke and systemic embolism.^21, 22^

Data from the Danish Stroke Registry^23^, however, revealed no significant differences in stroke severity and 1-year mortality between inappropriate and appropriate direct oral anticoagulant therapy. Notably, this study population was older, with an average age of 78.7 years. Similarly, a nationwide study focusing on elderly individuals (average age 82 years) demonstrated that over a one-year observation period, underdosed DOAC treatment did not significantly differ in effectiveness compared to recommended-dose DOAC treatment.^24^ Interestingly, the underdosed DOAC treatment was associated with a lower bleeding rate. A post hoc analysis of the ENGAGE AF-TIMI 48 randomized clinical trial found that in AF patients 80 years and older, those receiving edoxaban 30mg daily had lower major bleeding events compared to those receiving 60mg daily (in patients without dose-reduction criteria), without an increase in ischemic events.^25^ However, conflicting results exist. An Italian multicenter study of AF patients aged 80 and above found no significant differences in thromboembolic events, major bleeding events, and mortality rates among different dose subgroups. Notably, the underdosed group had a significantly lower survival rate compared to the appropriate dose group.^13^ A retrospective cohort study from Taiwan’s National Health Insurance Research Database (NHIRD) also showed that for AF patients aged 80 and above, low-dose DOACs were associated with higher risks of arterial and venous thrombosis, death, and composite outcomes compared to recommended-dose DOACs.^26^

Our study had several limitations. First, the retrospective study design may have led to bias and incompleteness in data collection. Second, the single-center study limited the generalizability of the results. Finally, the study struggled to comprehensively explain all the decision-making factors for physicians choosing specific dosages.

In conclusion, inappropriate DOAC dosing is very common among very elderly AF patients, particularly with underdosing. This situation has not significantly improved over the years. Therefore, there is an urgent need to promote patient-centered multidisciplinary management and shared decision-making models for AF, while also conducting high-quality prognostic studies.

## Abbreviations

AF: Atrial fibrillation
BMI: Body mass index
CrCl: creatinine clearance
DOAC: Direct oral anticoagulant
NVAF: non-valvular atrial fibrillation

## Supplementary material

**Supplementary Table S1.** Appropriate dosage levels for NOACs administered to patients aged ≥80 years with atrial fibrillation in China

**Supplementary Table S2.** Baseline patient characteristics between underdosed and Non-underdosed group

**Supplementary Table S3.** Baseline patient characteristics between overdosed and Non-overdosed group

## Data availability

Data are available upon reasonable request with the corresponding author.

## Author’ contribution

Ya-Tong Zhang: execution, analysis, writing the main manuscript text.

Jun-Peng Liu: funding acquisition, study design, supervision, editing, review.

Hong-Qiu Gu and Yu-Hao Wan: analysis, review.

Zi-Nan Zhao, Tian-qi Zhang, Yi-Fan Na, Min Dong and Min Zeng: investigation, review.

Ning Sun and Cheng Wu: investigation, data Curation, review.

Jie-Fu Yang: supervision, review.

